# Using serum metabolomics analysis to predict sub-clinical atherosclerosis in patients with SLE

**DOI:** 10.1101/2020.08.11.20172536

**Authors:** Leda Coelewij, Kirsty E Waddington, George A Robinson, Elvira Chocano, Thomas McDonnell, Filipa Farinha, Junjie Peng, Pierre Dönnes, Edward Smith, Sara Croca, Maura Griffin, Andrew Nicolaides, Anisur Rahman, Elizabeth C Jury, Ines Pineda-Torra

## Abstract

**Background:** Patients with systemic lupus erythematosus (SLE) have an increased risk of developing cardiovascular disease (CVD) and 30-40% have sub-clinical atherosclerosis on vascular ultrasound scanning. Standard measurements of serum lipids in clinical practice do not predict CVD risk in patients with SLE. We hypothesise that more detailed analysis of lipoprotein taxonomy could identify better predictors of CVD risk in SLE.

**Methods:** Eighty patients with SLE and no history of CVD underwent carotid and femoral ultrasound scans; 30 had atherosclerosis plaques (SLE-P) and 50 had no plaques (SLE-NP). Serum samples obtained at the time of the scan were analysed using a lipoprotein-focused metabolomics platform assessing 228 metabolites by nuclear magnetic resonance spectroscopy. Data was analysed using logistic regression and five binary classification models with 10-fold cross validation; decision tree, random forest, support vector machine and lasso (Least Absolute Shrinkage and Selection Operator) logistic regression with and without interactions.

**Results:** Univariate logistic regression identified four metabolites associated with the presence of sub-clinical plaque; three subclasses of very low density lipoprotein (VLDL) (percentage of free cholesterol in medium and large VLDL particles and percentage of phospholipids in chylomicrons and extremely large VLDL particles) and Leucine. Together with age, these metabolites were also within the top features identified by the lasso logistic regression (with and without interactions) and random forest machine learning models. Logistic regression with interactions differentiated between SLE-P and SLE-NP with greatest accuracy (0.800). Notably, percentage of free cholesterol in large VLDL particles and age were identified by all models as being important to differentiate between SLE-P and SLE-NP patients.

**Conclusion:** Serum metabolites are a promising biomarker for prediction of sub-clinical atherosclerosis development in SLE patients and could provide novel insight into mechanisms of early atherosclerosis development.

## Introduction

Systemic Lupus Erythematosus (SLE) is a multi-system autoimmune condition that predominantly affects women (women to men ratio of 9:1) with a prevalence of approximately 1 in 1000 in the UK [1]. Patients with SLE are at an increased risk of developing cardiovascular disease (CVD) compared to healthy people of the same age and gender [2]. In a large multinational study of 9547 patients with SLE, a quarter of deaths were attributed to CVD [3].There is a 5-10 fold increased risk of developing CVD in SLE patients compared to age and sex-matched controls reported in epidemiological data. Strikingly, the presence of SLE in women between the ages of 35-44 increases the risk of coronary artery disease (CAD) by 50 times [2].

The precise mechanism of this increased CVD risk is yet to be fully elucidated. Whilst traditional risk factors such as high blood pressure, diabetes and high cholesterol contribute to the increased risk, they fail to account for it fully [4]. The risk is likely to be multifactorial, resulting from a complex interplay of SLE-driven immunological dysfunction and traditional CVD risk factors [4].

Abnormalities in lipid profile are known to be a traditional risk factor for CVD and can also be affected by chronic inflammatory conditions such as SLE. Lipids are central to driving atherosclerosis, the main pathology underlying CVD. Various fractions of lipoproteins can be distinguished in blood on account of their size and density: High Density Lipoprotein (HDL), Low Density Lipoprotein (LDL) and Very Low Density Lipoprotein (VLDL). Dyslipidemias are present in over 70% of cases of premature coronary heart disease [5] and elevated plasma concentrations of LDL and VLDL can induce the development of atherosclerosis in the absence of other risk factors [6]. In contrast, HDL has anti-atherogenic properties that include macrophage cholesterol efflux, anti-oxidation and protection against thrombosis [7]. Conversely, McMahon and colleagues have demonstrated the existence of a subpopulation of pro-inflammatory HDL in patients with SLE and rheumatoid arthritis that promotes atherosclerosis and could be a biomarker for increased risk of developing CVD [8, 9]. Hypercholesterolaemia (defined as blood elevated total cholesterol (TC) and/or LDL-cholesterol (LDL-C) or non-HDL-cholesterol (HDL-C)) is found in 34-51% of patients with SLE [5] and is characterised by elevated levels of VLDL and triglycerides (TGs) and low HDL levels [10]. In addition, development of CVD in women with SLE has been found to be associated with smaller sub-fractions of LDL [11]. Studies in patients with traditional CVD have shown the ratio between serum apolipoprotein-B:apolipoprotein-A1 (ApoB:ApoA1), two lipid-associated proteins, is a more effective CVD predictor than routine cholesterol measurements, a higher ratio is associated with increased cardiovascular risk [12-16], the role of this ratio in the prediction of SLE cardiovascular disease is still being assessed. However overall, dyslidaemia detected in routine lipid screens available in clinical practice, fail to account fully for the increased risk of CVD in patients with SLE [10]. Many SLE patients with normal lipid profiles on standard assays also go on to have CVD. Therefore, more sensitive and specific lipid profiles need to be delineated in order to identify high CVD risk patients in SLE cohorts.

Here we used a Nuclear Magnetic Resonance (NMR) Spectroscopy metabolomics platform [17] and machine learning (ML) analysis to assess the association of lipoprotein subclasses and lipid content and other serum metabolites with the presence of sub-clinical atherosclerotic plaque in patients with SLE.

## Materials and Methods

### Patient cohort

Serum samples were collected from 80 non-fasting patients with SLE attending a rheumatology clinic at University College London Hospital (UCLH) and fulfilling the American College of Rheumatology (ACR) classification criteria for lupus (1997)[18]. Patients had no previous history of CVD (defined as coronary artery disease, stroke, or myocardial infarction with confirmatory evidence from blood tests and/or imaging) and were scanned by vascular ultrasound between 2011-2013 [19].

Carotid and femoral ultrasound scans were performed to identify the presence of plaques. Demographic and clinical information were recorded at the time of scan/blood sampling, including sex, age, ethnicity, mean blood pressure (BP, average between systolic and diastolic BP), treatment (including hydroxychloroquine (HCQ), statins, ACE inhibitor, immunosuppressives, rituximab, prednisolone (and dose), and aspirin), and disease activity assessed by the global British Isles Lupus Assessment Group (BILAG) index [20] (Table 1). In total, four patients were not on any treatment at the time of the scan. All patients gave informed written consent and the study was approved by the combined UCL/UCLH Research Ethics Committee (Reference 06/Q0505/79).

**Table 1.**
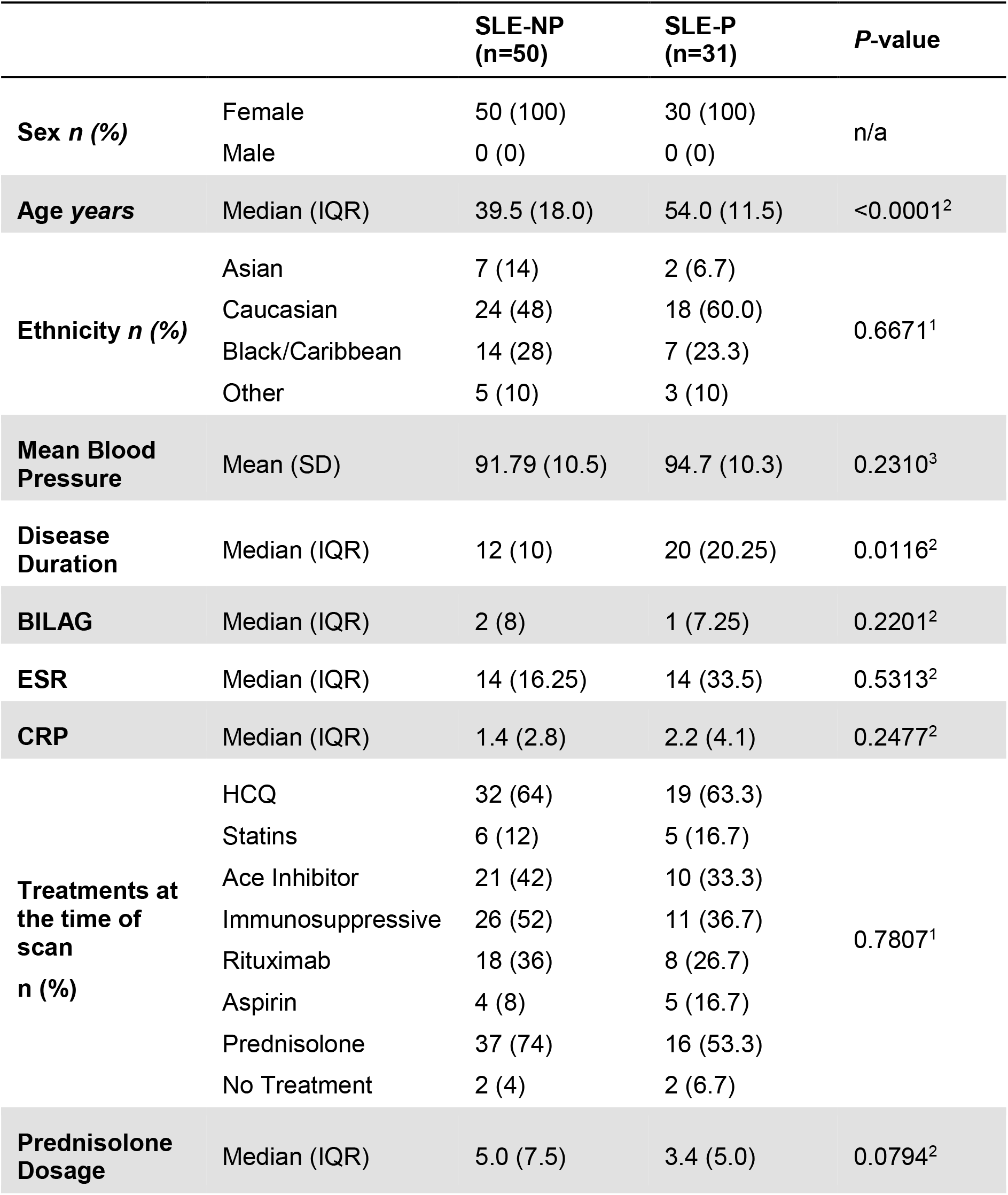
Cohort Characteristics. Demographic and clinical characteristics at the time of the first scan were compared between SLE patients with plaque (SLE-P) or those with no plaque (SLE-NP). Statistical comparisons were made using 1chi-squared or 3Mann-Whitney U. Abbreviations: BILAG – British Isles Lupus Assessment Group disease activity score, CRP – C reactive protein, ESR – erythrocyte sedimentation rate, IQR – interquartile range.

|  |  | **SLE-NP**<br>**(n=50)** | **SLE-P**<br>**(n=31)** | **P-value** |
| --- | --- | --- | --- | --- |
| **Sex n (%)** | Female | 50 (100) | 30 (100) | n/a |
|  | Male | 0 (0) | 0 (0) |  |
| **Age years** | Median (IQR) | 39.5 (18.0) | 54.0 (11.5) | <0.0001<sup>2</sup> |
| **Ethnicity n (%)** | Asian | 7 (14) | 2 (6.7) | 0.6671<sup>1</sup> |
|  | Caucasian | 24 (48) | 18 (60.0) |  |
|  | Black/Caribbean | 14 (28) | 7 (23.3) |  |
|  | Other | 5 (10) | 3 (10) |  |
| **Mean Blood Pressure** | Mean (SD) | 91.79 (10.5) | 94.7 (10.3) | 0.2310<sup>3</sup> |
| **Disease Duration** | Median (IQR) | 12 (10) | 20 (20.25) | 0.0116<sup>2</sup> |
| **BILAG** | Median (IQR) | 2 (8) | 1 (7.25) | 0.2201<sup>2</sup> |
| **ESR** | Median (IQR) | 14 (16.25) | 14 (33.5) | 0.5313<sup>2</sup> |
| **CRP** | Median (IQR) | 1.4 (2.8) | 2.2 (4.1) | 0.2477<sup>2</sup> |
| **Treatments at the time of scan n (%)** | HCQ | 32 (64) | 19 (63.3) | 0.7807<sup>1</sup> |
|  | Statins | 6 (12) | 5 (16.7) |  |
|  | Ace Inhibitor | 21 (42) | 10 (33.3) |  |
|  | Immunosuppressive | 26 (52) | 11 (36.7) |  |
|  | Rituximab | 18 (36) | 8 (26.7) |  |
|  | Aspirin | 4 (8) | 5 (16.7) |  |
|  | Prednisolone | 37 (74) | 16 (53.3) |  |
|  | No Treatment | 2 (4) | 2 (6.7) |  |
| **Prednisolone Dosage** | Median (IQR) | 5.0 (7.5) | 3.4 (5.0) | 0.0794<sup>2</sup> |

### Plaque detection

Scans were performed by an experienced vascular scientist using strict scanning protocols and established equipment settings (Griffin et al., 2009; Nicolaides et al., 2010). Vascular ultrasound scans of the common carotid artery, carotid bulb, carotid bifurcation, common femoral artery and femoral bifurcation were performed bilaterally using the Philips IU22 ultrasound computer and the L9-3 MHz probe. IMT measurements were performed using QLAB Advanced Quantification Software® version 7.1 (Philips Ultrasound, Bothell, USA). Plaque was defined as “a focal thickening >1.2 mm that encroaches into the arterial lumen as measured from the media-adventitia interface to the lumen interface” [21]. Patients having at least one region fulfilling this description were included in the group with plaque (SLE-P).

Ultrasound images were stored as DICOM files and analysed using Carotid Plaque Texture Analysis software for ultrasonic arterial wall and atherosclerotic plaque measurements (LifeQ Medical Ltd – www.lifeqmedical.com). Total plaque area (TPA) was defined as the sum of the cross-sectional areas of all plaques seen in longitudinal images (plaque area in mm2). Grey Scale Median (GSM): Plaque echogenicity was expressed numerically by GSM value [22], a measure of plaque stability and lipid content. Lower GSM values signify more echolucent plaque associated with lipid and inflammatory cell content; whereas plaque with high GSM scores are more stable and less echolucent [23]. Images were normalized using linear scaling with two reference points blood (grey scale=0) and adventitia (grey scale=190).

**Table 2.** Comparison of predictive model performance. Performance statistics for 5 predictive models based on serum metabolites at the time of the first scan. The models used were logistic regression (LR) with and without interactions (i), support vector machine (SVM), random forest (RF), and decision tree (Tree). The classification accuracy (CA) represents the proportion of correctly identified cases, in contrast to specificity, which is the true negative rate. F1 is the weighted average of the precision and recall (see Methods). Statistics are rounded to 3 decimal places.

| Model | F1 | Precision | Recall | Specificity | CA | AUC |
| --- | --- | --- | --- | --- | --- | --- |
| LR | 0.630 | 0.708 | 0.567 | 0.860 | 0.750 | 0.802 |
| LR + I | 0.714 | 0.769 | 0.667 | 0.880 | 0.800 | 0.812 |
| SVM | 0.480 | 0.600 | 0.400 | 0.840 | 0.675 | 0.707 |
| RF | 0.542 | 0.722 | 0.433 | 0.900 | 0.725 | 0.779 |
| Tree | 0.464 | 0.500 | 0.433 | 0.740 | 0.625 | 0.630 |

### Serum metabolomics analysis

Measures of 228 serum biomarkers were acquired with an established nuclear magnetic resonance (NMR)-spectroscopy platform (Nightingale Health)[24, 25]. These included both absolute concentrations (mmol/L), ratios, and percentages (%) of lipoprotein composition. Serum lipids measured included apolipoproteins (Apo) and VLDL, LDL, intermediate density lipoprotein (IDL) and HDL particles of different sizes ranging from chylomicrons and extremely large (XXL), very large (XL), large (L), medium (M), small (S) and very small (XS). Lipids within each lipoprotein subclass included –total lipid (L), phospholipids (PL), total cholesterol (C), cholesterol esters (CE), free cholesterol (FC) and triglycerides (TG). Distribution of these lipids was expressed as a ratio or percentage of total lipid content for each lipoprotein subclass (for list of metabolites see Supplementary Table 1).

**Table 3.** Predictors for lasso logistic regression with interactions. List of features selected by the lasso regressions with interactions. Interacting features are separated by a colon (:). Interactions are listed in order of the absolute size of the beta coefficient, where the top ten metabolite interactions will have the largest effect on the model classification. Where predictors are categorical (ethnicity) the specific category is shown in brackets. Abbreviations: BP – blood pressure, HCQ – hydroxychloroquine

| Features Included | Beta Coefficient |
| --- | --- |
| Intercept | -1.17411 |
| S-LDL-CE : Rituximab | -0.68492 |
| S-HDL-FC : XXL-VLDL-CE_% | -0.49192 |
| Gly : His | -0.46405 |
| Statins : ACE Inhibitor | -0.37603 |
| Lac : Cit | -0.37161 |
| HCQ : ACE Inhibitor | -0.23705 |
| XL-HDL-TG : Glol | -0.15531 |
| IDL-CE : Glol | -0.12971 |
| His : XS-VLDL-CE_% | -0.09162 |
| L-VLDL-TG : Lac | -0.08858 |
| Glol : XXL-VLDL-CE_% | -0.07212 |
| Gly : XS-VLDL-CE_% | -0.05998 |
| Glol : His | -0.03934 |
| DHA/FA : Age | 0.019318 |
| DHA/FA : Tyr | 0.019945 |
| L-HDL-PL_% : Age | 0.022831 |
| L-VLDL-TG : HCQ | 0.034663 |
| Ethnicity (Caucasian) : Disease Duration | 0.035969 |
| XXL-VLDL-PL_% : Mean Blood Pressure | 0.076793 |
| DHA : Aspirin | 0.089295 |
| DHA : HCQ | 0.096398 |
| HCQ : Prednisolone Dose | 0.129875 |
| Age : Mean Blood Pressure | 0.130915 |
| SFA/FA : M-VLDL-FC_% | 0.193219 |
| Tyr : Disease Duration | 0.194325 |
| M-VLDL-FC_% : L-LDL-FC_% | 0.239355 |
| HCQ1 : Rituximab | 0.282773 |
| IDL-TG_% : S-LDL-C_% | 0.292232 |
| SM : Tyr | 0.333199 |
| Leu : Mean Blood Pressure | 0.401734 |
| Gln : Ace | 0.464306 |
| SM : Leu | 0.597411 |
| Ethnicity (Black / Caribbean) : HCQ | 0.755513 |
| L-VLDL-FC_% : Age | 1.056669 |
| IDL-TG_% : Age | 1.133333 |

### Logistic regression

To assess the association of plaque with NMR metabolomic biomarker data, univariate logistic regressions were performed for each individual serum metabolite, adjusted for ethnicity, age, mean blood pressure, global BILAG score, and treatments at the time of the scan. Results were visualized in a forest plot using R package forestplotNMR [26].

### Predictive models

The data analysis pipeline used in shown in Figure 1. RStudio (The R Foundation, Vienna, Austria) (R Core Team, 2019) and Orange 3.24.1 (Bioinformatics Lab, University of Ljubljana, Slovenia) [27] were used for machine learning analysis as we have shown previously [28]. Five different supervised learning algorithms were implemented: support vector machine (SVM), logistic regression with and without interactions, decision trees, and random forest classification. The outcome of the learning algorithms was to predict whether an SLE patient is likely to develop plaque. Predictive models were generated from metabolite concentrations at the time of the scan.

**Figure 1.**
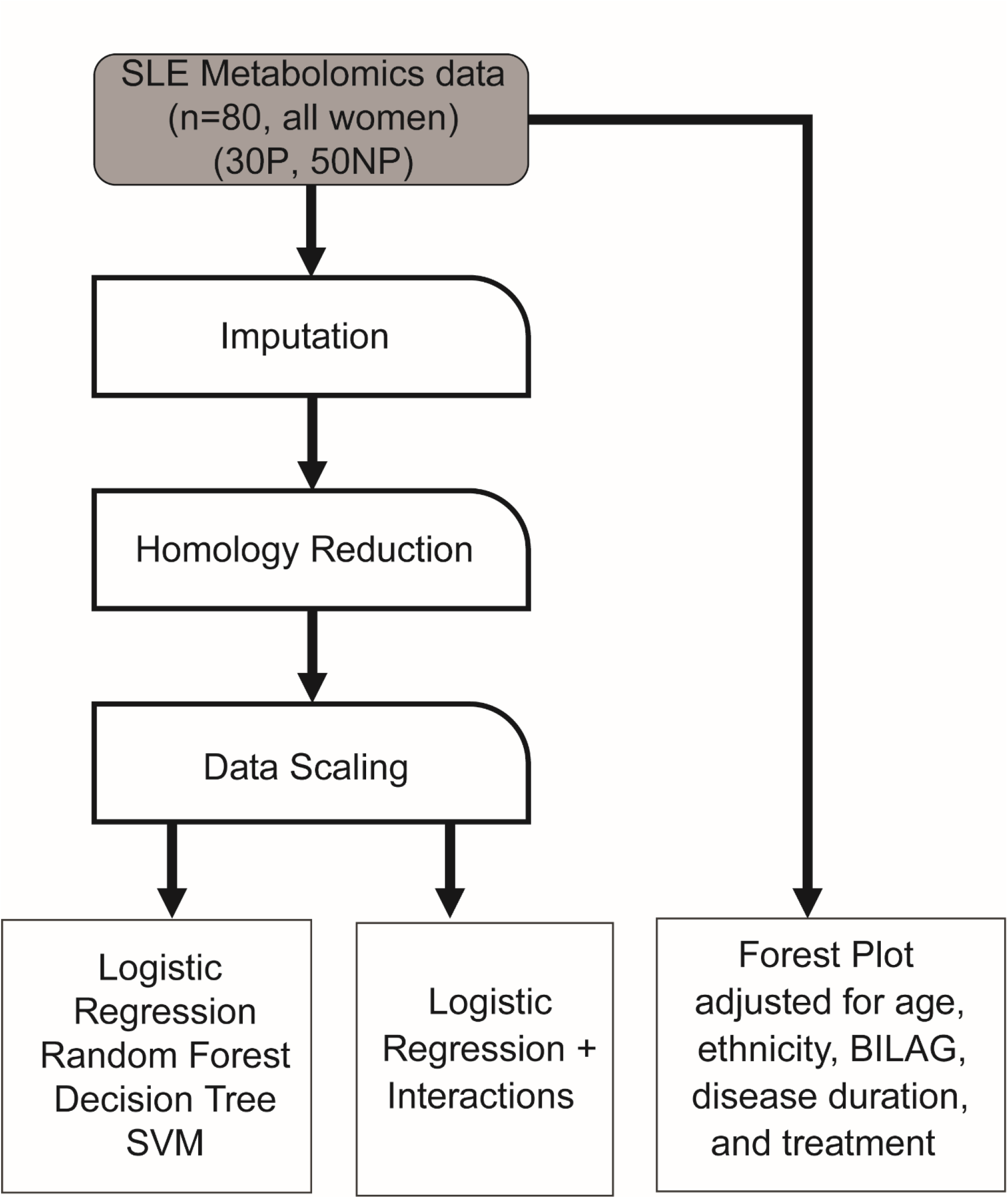
Data analysis workflow. Flow chart depicting the data cleaning and processing steps taken prior data analysis using machine learning algorithms. Abbreviations: SVM – support vector machine | P – plaque | NP – no plaque, BILAG – British Isles Lupus Assessment Group disease activity score.

*Missing data:* Features with >10% missing data were excluded (pyruvate). Remaining missing values (n=5) were imputed using k-nearest neighbors with k = 5.

*Homology reduction:* To reduce homology, if two features had a correlation co-efficient >0.95 then the feature with the greatest mean absolute correlation with the remaining features was removed (Supplementary Data File 1). This left 124 metabolites.

*Data scaling:* Metabolite concentrations were centered on the mean and scaled to the standard deviation.

*Predictors:* The independent variables included in the models were the homology reduced dataset (124 metabolites), as well as the cohort information (age, ethnicity, mean blood pressure, disease duration, BILAG, treatments at the time of first scan). Sex was not considered as all participants were female. Full lists of the predictors contributing to each model are included in Supplementary Data File 2.

*Support vector machine (SVM):* A supervised classification method which creates a hyperplane to optimally separate data into two classes [29]. As this data set was not linearly separable, the radial basis function kernel was used. Values for C, epsilon, and gamma were tuned using the R Package e1071 [30]. The parameters were set to C = 4.0, epsilon = 0.1, gamma = 0.01.

*Decision tree:* A supervised machine learning method which classifies incidents according to their features. Decision trees are built using forms of impurity measures, such as information gain and entropy [31]. To prevent overfitting, decision trees were limited to a depth of 4 and subsets of 5 or less were not split further.

*Random Forest (RF):* A statistical classifier (machine-learning algorithm) that assigns observations into classes by creating a set of decision trees, or ‘forest’. Importance of each feature was quantified by the Gini index, which represents the total variance across the two classes, the purity of each node and the quality of each split. The values for mtry and ntree were tuned using the R Package [32]. The parameters were set to mtry=10, ntree=10,000.

*Logistic regression with/without interactions (LR/LR+I):* The least absolute shrinkage and selection operator (lasso) method uses the absolute value of the co-efficient as a penalty to shrink less important features to zero. The strength of shrinkage is determined by tuning the regularization variable lambda (λ). Ln(λ) was optimized using the R package [32] and set to lambda=0.059 and lambda=0.065 for logistic regression with and without interaction respectively.

*Model performance:* Ten-fold cross-validation was used to evaluate model performance. Validation was performed in Orange for the decision tree and in R for all other models, using a balanced splitting. The following performance metrics were calculated from the confusion matrices: (i) F1 score - a weighted average of precision (positive predictive value) and recall (sensitivity), (ii) specificity - the true negative rate, and (iii) classification accuracy (CA) - the proportion of correctly classified cases.

### Partial Least Squares Discriminant Analysis (sPLS-DA)

A sPLS-DA is a supervised clustering machine-learning approach that combines parameter selection and classification into one operation. This analysis was performed using the features selected for the lasso LR+I model (Table 3).

### Statistical testing

Statistical tests were performed in Microsoft Excel and GraphPad Prism version 8.3.0 for Windows (GraphPad Software, San Diego, USA). Data was assessed for normality and analysed with parametric or non-parametric tests as appropriate. Details of statistical tests are given in the figure legends. P-values < 0.05 were considered statistically significant.

## Results

### Serum metabolites can predict the presence of pre-clinical atherosclerotic plaque in SLE patients

Metabolites were quantified in serum from SLE patients classed as having plaque(s) (SLE-P) or not having plaque(s) (SLE-NP) using vascular ultrasound (Table 1 and Supplementary Table 1). Several analysis strategies were applied to identify metabolites associated with sub-clinical plaque development (Figure 1). All models were adjusted for ethnicity, age, mean blood pressure (BP), disease duration, disease activity (global BILAG score), and treatment at the time of the scan. Firstly, univariate logistic regressions identified four metabolites which differentiated between SLE-P and SLE-NP patients; Leucine, M-VLDL-FC_%, L-VLDL-FC_% and XXL-VLDL-PL_% which were all increased in serum from SLE-P compared to SLE-NP patients (Figure 2A, Supplementary Data File 3 and 4). Of note, these metabolites can be significantly affected by treatment with statins [33, 34] and pro-protein convertase subtilisin/kexin type 9 (PCSK9) inhibitors [35], or body mass index (BMI) [36], suggesting that abnormal serum lipid metabolite profiles in SLE-P patients, could be modified using available therapies or interventions (Figure 2A asterisks).

**Figure 2.**
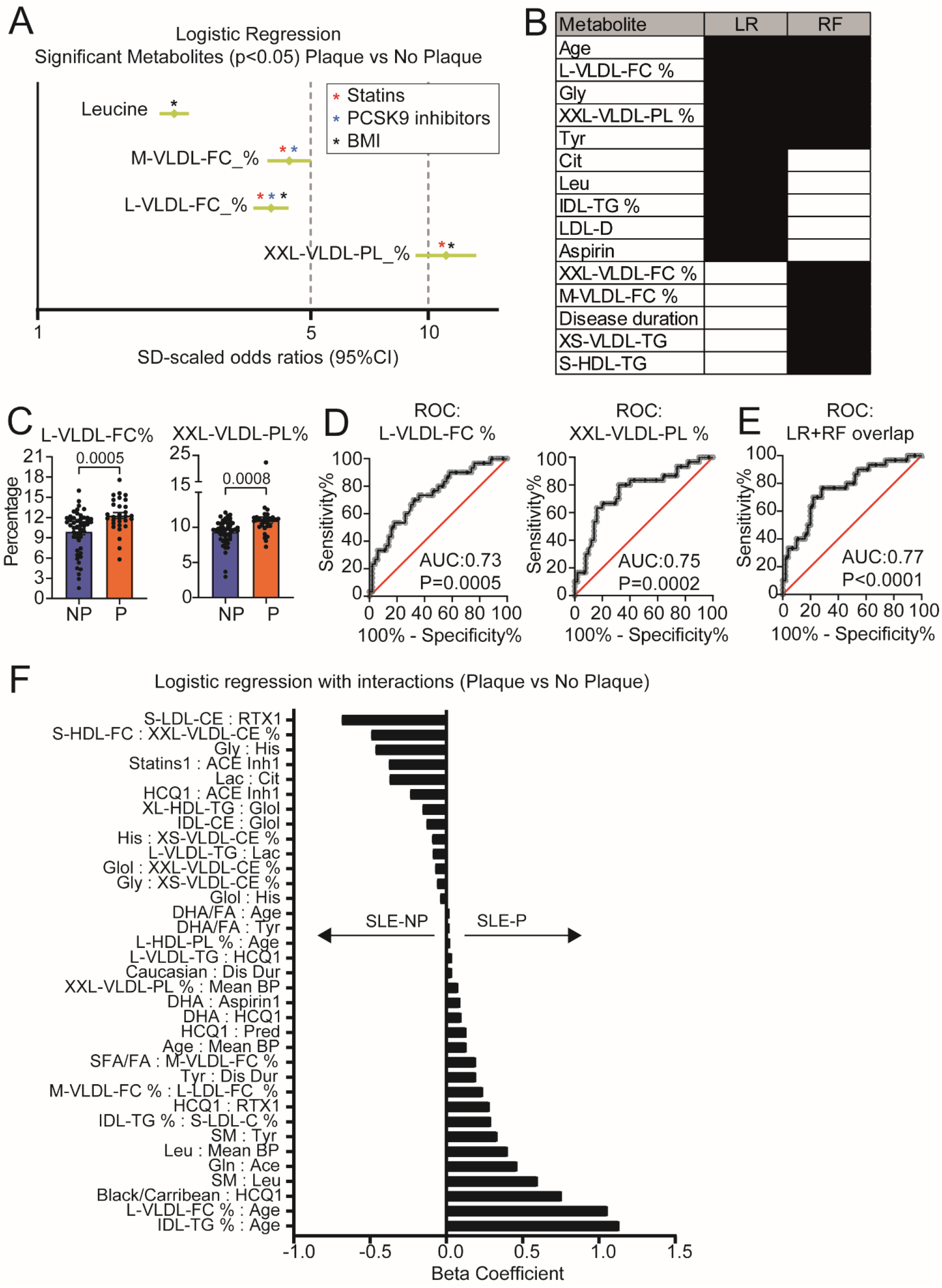
Identification of important metabolites separating patients with SLE-P from SLE-NP. (**A)** Forest plot depicting statistically significant individual logistic regression results of metabolites in SLE-P vs SLE-NP. Results given in Odds Ratio (95% confidence intervals). Logistic regressions were adjusted for age, ethnicity, mean blood pressure, global BILAG, disease duration, and treatments at the time of scan. Coloured asterisks denote metabolite has previously been shown to be modified by statins, PCSK9 inhibitors or BMI [33-36]. (**B**) Best performing models were determined based on performance statistics (Table 2). The top ten features of these LR and RF models are listed to identify common features. (**C-D**) Metabolites which were significant in the individual logistic regression and featured in the top ten of the LR and RF models were further analysed using **(C)** bar charts showing mean, and p value (see Supplementary Figure 2A) and **(D)** ROC plots (see Supplementary Figure 2B). **(E)** Metabolites which featured in the top ten of the LR and RF model were combined and analysed in a single ROC plot (L-VLDL-FC%, Glycine, XXL-VLDL-PL%, Tyrosine). (**F**) Beta coefficients from the logistic regressions with interactions are plotted for the SLE-P vs SLE-NP analysis. The sign indicates the direction of the effect of the predictor; a positive sign indicates an increased likelihood of a SLE-P prediction, while a negative sign indicates an increased likelihood of a SLE-NP prediction.

Next, five supervised ML models were developed and validated to predict the presence of plaque in SLE patients; LR, LR+I, SVM, RF and Decision tree. Since many of the metabolites measured were biologically interdependent, and therefore highly correlated, homology reduction was applied (see methods and Supplementary Data File 1). The models were built using the homology reduced dataset (124 metabolites) and patient information (age, ethnicity, mean BP, disease duration, global BILAG index, treatments at the time of first scan). Sex was not considered as all participants were female (Supplementary Data File 2 for full lists of the predictors contributing to each model).

The top three models, according to classification accuracy, specificity, and F1 scores, were LR, LR+I, and RF (Table 2). Performance metrics were based on predictions of the models, summarised in confusion matrices (Supplementary Figure 1). LR and LR+I had a similar performance, correctly classifying 75% and 80% of SLE-P patients respectively. The RF model had the best specificity, identifying 45 out of 50 (90%) SLE-NP patients correctly. The three models were further investigated to identify the top features in predicting plaque formation in SLE (Figure 2B-E). Four metabolites (XXL-VLDL-PL_%, L-VLDL-FC_%, glycine and tyrosine) and patient age were identified in both LR and RF models as important predictors for SLE-P (Figure 2B). Metabolites which were significant in individual logistic regressions and also featured in the top ten metabolites of the LR and RF model, were further investigated for differences between SLE-P and SLE-NP patients (Figure 2C and Supplementary Figure 2A). Receiver operating characteristic curve (ROC) of these metabolites showed an area under the curve (AUC) of 0.6810 (XXL-VLDL-PL %), 0.7523 (L-VLDL-FC %), 0.7337 (glycine), and 0.6300 (tyrosine) (Figure 2D and Supplementary Figure 2B), demonstrating a potential predictive ability of these metabolites independent of other clinical features and metabolites. When the four shared metabolites between LR and RF models were combined, the ROC analysis showed an AUC of 0.7667 (Figure 2E), demonstrating a stronger potential predictive ability of the metabolites grouped together.

The LR+I model included interaction of each metabolite with all other metabolites and clinical features (assessing over 15,000 possible features) and was therefore assessed separately from the other models. This model performed best overall, with a classification accuracy of 0.800 (Table 2). Using the lasso method of shrinkage and selection, only 35 interactions were identified as important and given a beta coefficient to describe the effect size and direction on the model (Figure 2F, Table 3). Features with larger beta coefficients had the greatest effect on the classification. Notably, L-VLDL-FC_%: age and IDL-TG_%:age, Black/Carribean ethnicity:hydroxycloroquine treatment and sphingomyelin:leucine were significantly associated with plaque; while S-LDL-CE:rituximab treatment, S-HDL-FC:XXL-VLDL-CE_%, glycine:histidine and statins:ACE inhibitor treatment were associated with no-plaque.

### Metabolite interactions can differentiate between SLE-P and SLE-NP patients

Using the metabolite interaction features that were selected for the LR+I model (Table 3, Figure 2F), a sPLS-DA was performed to rank and validate the metabolite features by their distribution in patients with SLE-P and SLE-NP. By assessing the overall estimation error rate in ten-fold cross-validation, models with 4 components and a subset of 28 metabolite features were chosen for optimal model performance (Figure 3A). This analysis identified a significant separation between SLE-P and SLE-NP patients by plotting principal component (PC)-2 against PC-1 (Figure 3B). The 28 selected metabolite interaction features were ranked by discriminating capability (Figure 3C, 3D). The two highest weighted features were L-VLDL-FC_%:Age and IDL-TG_%:Age, which were also the highest ranked features in the LR+I model (Figure 2F). L-VLDL-FC_% and age were both also featured as one of the top ten features shared by LR and RF (Figure 2B) and were differentially expressed between SLE-P and SLE-NP patients (Figure 2A, Table 1) suggesting an influential role of both features in SLE-P patients.

### Differential metabolites correlated with clinical features of SLE-P patients

Finally, the top ten metabolites from LR, RF, and all metabolite interactions included in the LR+I models were correlated with clinical and plaque features (Figure 4, Supplementary Table 2). Significant correlations include GSM (grey scale median, measure of plaque stability and lipid content) correlated positively with S-HDL-TG, XS-VLDL-TG, XL-HDL-TG:Glycerol and IDL-CE:Glycerol and negatively with LDL-D, His:XS-VLDL-CE_% and Glycine:XS-VLDL-CE_%; Plaque number and plaque thickness negatively correlated with His:XS-VLDL-CE_%; TPA (total plaque area) positively correlated with L-VLDL-TG:Lactate; and disease activity (BILAG) positively correlated with M-VLDL-FC_% and M-VLDL-FC_%:L-LDL-FC_% and negatively with DHA-FA:age. The strongest correlation was between disease duration and Tyrosine:disease duration.

**Figure 3.**
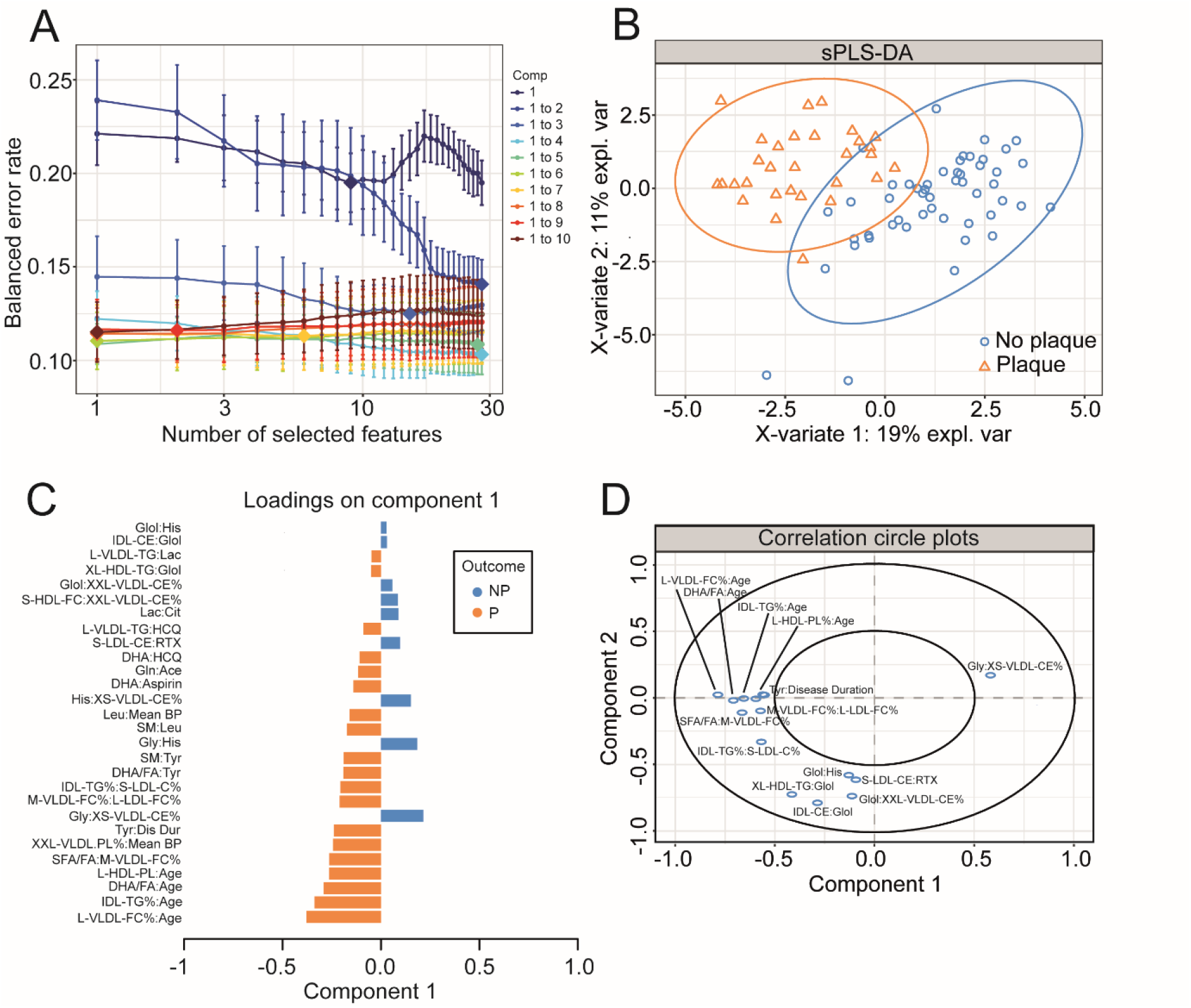
Partial least squares discriminate analysis validated metabolites identified by logistic regression with interactions to predict SLE-P. (**A**) Model optimisation – model with different components and features kept in the analysis were analysed, with each colour representing a different number of components (Comp), number of features kept in the analysis on the x-axis, and the overall error on the y-axis. (**B**) sPLS-DA plot to validate top hits from the logistic regression with interactions. sPLS-DA is a supervised clustering method which separates SLE-P from SLE-NP. (**C**) Features included in the sPLS-DA plotted with their factor loading value. (**D**) Visualisation of the weighting and correlation of each metabolite in component 1 and 2 on the sPLS-DA model.

**Figure 4.**
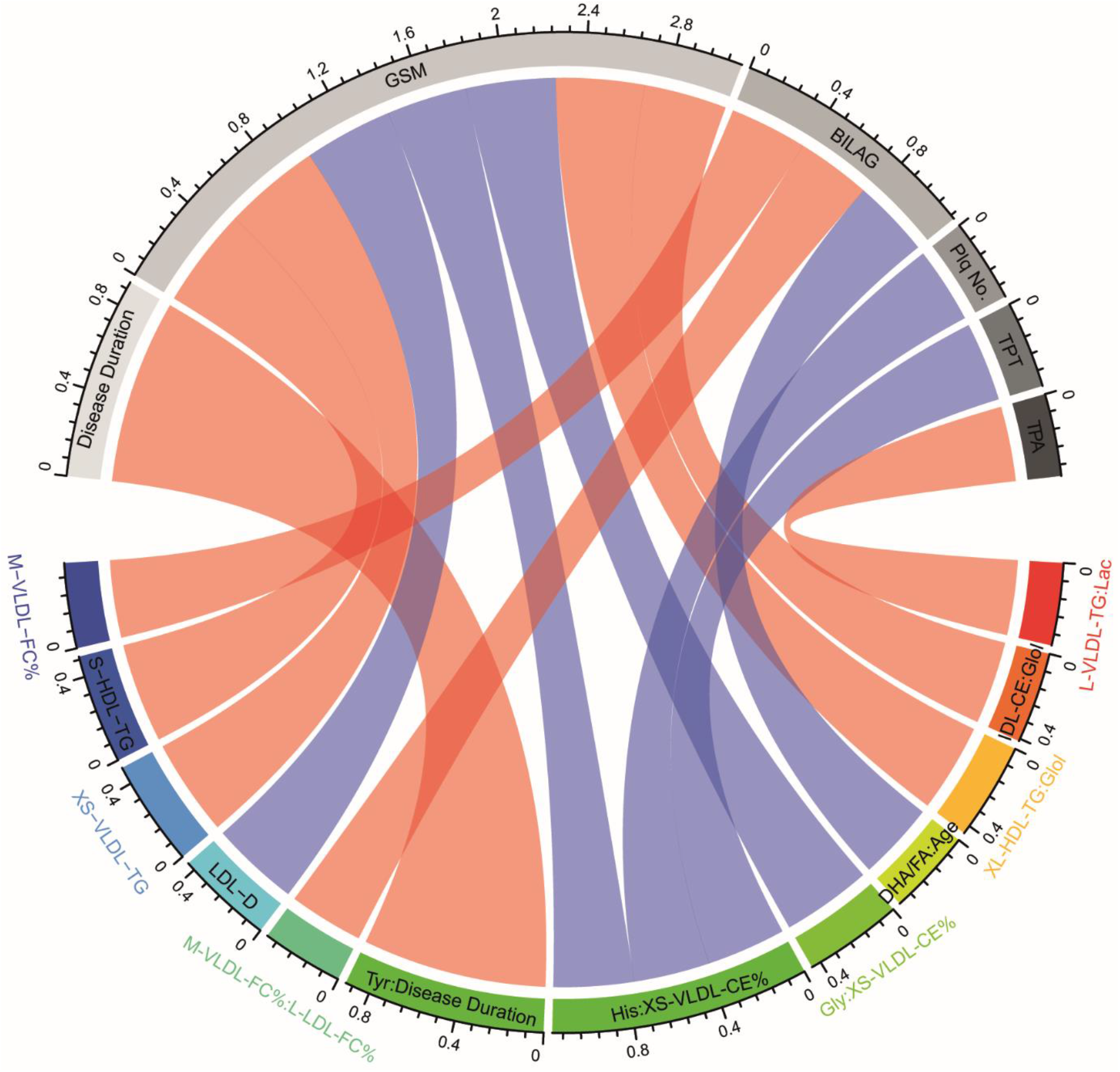
Correlations of significant metabolites with SLE clinical markers. Correlations between metabolites and patient clinical characteristics. Pearson’s product moment correlation coefficients are represented as connecting lines between the clinical characteristic section (grey) and metabolite section (rainbow). Only correlations with p value below 0.05 are shown. Red line=positive correlation and blue line=negative correlation. Width of lines represents the value of correlation coefficients (measured with scale). GSM – grey scale median, BILAG – British Isles Lupus Group Index disease activity score, Plq No – plaque number, TPT – total plaque thickness, TPA – Total Plaque area. (see Supplementary Table 1 for metabolite abbreviations.

## Discussion

CVD risk in SLE patients is an important cause of mortality in a cohort of mostly female and relatively young patients compared to the general population. Despite the long-established link between SLE and increased CVD risk [2, 4, 37] SLE specialists still lack accurate methods of predicting the risk in an individual patient. Traditional cardiovascular risk factors encapsulated in the Framingham equations underestimate the true risk in patients with SLE and fail to predict which patients will have cardiovascular events [38, 39]. SLE specialists therefore have no means currently of stratifying patients at high risk.

This study used serum metabolomics incorporating detailed lipoprotein subclass evaluation to differentiate between patients with SLE with and without confirmed sub-clinical atherosclerosis. Analysis using machine learning models identified an association between multiple VLDL subsets, the branched-chain amino acid Leucine and clinical features including age with the presence of sub-clinical plaque, suggesting that more detailed lipoprotein and metabolomic measurements together with demographic information could help to better predict those patients at greatest CVD risk.

VLDL subclasses featured predominantly in all the analysis models used suggesting its potential importance in predicting which patients with SLE go on to develop atherosclerosis. VLDL is known to be associated with increased CVD risk. It is the main carrier of triglycerides, which are an independent risk factor for CVD [40], and VLDL particle concentrations have been positively associated with the risk of myocardial infarction [41]. In the JUPITER trial of CVD risk in nearly 12,000 patients, risk among placebo‐allocated participants was associated with total VLDL particles, as well as apolipoprotein B, total cholesterol, and TGs [42]. Furthermore, pharmacological lowering of apolipoprotein(Apo)-B–containing lipoproteins, including VLDL, earlier in life is proposed to eliminate high risk of atherosclerotic-CVD in individuals with image-documented sub-clinical atherosclerosis, such as the SLE women in this study [43]. Mechanistically, larger, typically TG-rich, apoplipoprotien-(apo) B-containing lipoproteins such as VLDL, may have difficulty leaving the intima because of their larger size or because they get entrapped by components in the subendothelial space [44]. Here, these lipoproteins undergo enzymatic modifications that accelerate accumulation and promote aggregation, which is influenced by lipoprotein quantity and composition [45]. Notably, the larger TG and cholesterol-rich lipoprotiens appear to be more potent than LDL, the most common atherogenic lipoprotein, for provoking greater maladaptive immune activation [46]. This supports our previous work showing that VLDL from SLE-P patients could influence the phenotype of *i*NKT cells and monocytes [47].

Previous metabolomics studies in patients with SLE have not focused on cardiovascular risk but rather compared metabolomics profiles between healthy donors and SLE patients [48]. One study used mass spectroscopy rather than NMR to compare 20 patients with SLE and 9 healthy controls, identifying >100 differentially expressed metabolites, but did not assess lipoprotein particles and only one patient had CVD [49]. Another study using NMR, identified raised VLDL and LDL and reduced HDL in patients with SLE [50]. However, they did not report on lipoprotein subclasses and vascular scanning was not performed. Guleria *et al* showed that metabolomics could be used to compare different clinical subgroups within a cohort of lupus patients [51], as we have done here for SLE-P versus SLE-NP patients. Another NMR metabolomics study compared patients with and without lupus nephritis, and healthy controls. Compared to healthy controls, this study reported lower VLDL and LDL in patients with SLE, though higher in the nephritis patients [50]. The study was done in India so ethnicity and lifestyle factors such as diet may have played a role in these results, which do not support other reports.

We also found associations between metabolites and scanning outcomes. Interestingly, TGs in VLDL showed significant correlations with TPA and GSM. Measurements of TPA and echolucency have been shown to have good predictive value for coronary artery disease in women [52], and thus pertinent in our all-female SLE cohort.

In addition to lipoproteins, Leucine, Glycine and Tyrosine were also identified by the ML models to be important predictors of SLE-P. Leucine is a branched chain essential amino acid with a potential important role in regulating protein, glucose and lipid metabolism, in part via activation of the mTOR (mammalian target of rapamycin) protein kinase and promotion of leptin synthesis [53]. Studies in mice have shown that Leucine supplementation improved diet-induced obesity, insulin resistance and atherosclerosis outcomes [54, 55], the increased Leucine in this study and could be associated with an early response to sub-clinical plaque development as we have shown previously[47]. Glycine, shown here to be reduced in SLE-P, is also reported have a potential anti-inflammatory role via reducing nuclear factor-kappa B (NF-KB) activation in vascular endothelial cells [56]. Plasma Glycine levels are inversely correlated with acute myocardial infarction in patients undergoing coronary angiography [57]. Tyrosine, in the form of 3-nitrotyrosine, is associated with oxidised HDL in the human artery wall and circulation in atherosclerosis, and may promote atherogenesis [58]. Further work is needed to confirm these findings and to understand fully the complex role of these metabolites in atherogenesis in SLE.

In conclusion, the interrogation of lipid subclasses may hold the key to providing insights on how to stratify CVD risk in SLE. Analysis of lipoproteins using NMR spectroscopy is of particular interest given the high throughput metabolomics analysis is rapid, can be carried out on serum, gives a larger amount of information from each sample and is potentially cost-effective [59] depending on how many high-risk patients are identified and how the risk is managed. It will be important to validate our results in larger, multi-centre cohorts. It is likely that a composite score of metabolomics with conventional risk factors may be the best way to assess CVD risk in patients with SLE, as in the type of extended risk model proposed by Wurtz *et al* [60].

## Data Availability

Metabolomic data will be made available in an open access repository upon acceptance for publication in a peer reviewed journal

## Contributors

IPT and ECJ conceived the study, secured the funding and supervised all aspects of the work. Acquiring data; ES, KEW, SC. Recruiting patients; AR, SC, ES. Performing vascular scans: AN, MG. Analyzing data; LC, EC, GAR, KEW, JP, ECJ, IPT, PD, TMcD, FF. Preparing figures: LC, GAR, JP. Writing the manuscript; LC, ECJ, KEW, AR, IPT.

## Acknowledgments

LC is supported by UCL & Birkbeck MRC Doctoral Training Programme. GAR was supported by a PhD studentship from Lupus UK and The Rosetrees Trust (M409). KEW was funded by a British Heart Foundation PhD Studentship (FS/13/59/30649). JP is supported by Versus Arthritis grant reference 21226. This work was also supported by NIHR UCLH Biomedical Research Centre grant reference BRC531/III/IPT/101350 (IPT/ECJ).

## Competing interests

The authors have declared that no conflict of interest exists.

## Notes

### Competing Interest Statement

The authors have declared no competing interest.

### Author Declarations

Combined University College London/University College London Hospitals Research Ethics Committee (Reference 06/Q0505/79).

